# Immunogenicity of JN.1- and KP.2-Encoding mRNA COVID-19 Vaccines Against JN.1 Subvariants in Adult Participants

**DOI:** 10.1101/2025.05.02.25325954

**Authors:** Amparo L. Figueroa, Bethany Girard, Darin K Edwards, Arshan Nasir, Kimball Johnson, Steven Hack, Xin Cao, Elizabeth de Windt, Veronica Urdaneta, Frances Priddy, Rituparna Das, David C Montefiori, Spyros Chalkias

**Affiliations:** Moderna, Inc., Cambridge, Massachusetts 02142, USA; CenExel, Decatur, Georgia 30030, USA; Duke University Medical Center, Durham, North Carolina 27710, USA

**Keywords:** immunogenicity, SARS-CoV-2, KP.2 vaccine, JN.1 vaccine, mRNA-1273

## Abstract

In this ongoing, open-label phase 3b/4 study, JN.1- and KP.2-encoding monovalent mRNA-1273 vaccines elicited robust neutralizing antibody responses against vaccine-matched variants and cross-neutralized currently circulating JN.1 subvariants (KP.3.1.1, XEC, LP.8.1) in adults with previous COVID-19 mRNA vaccination, with reduced cross-neutralization observed against all subvariants tested. No safety concerns were identified.

## Background

Rapid spread and diversification of the SARS-CoV-2 Omicron lineage led to the JN.1 variant, which became globally dominant by early 2024 [1] and continues to diversify, with KP.2 having circulated previously, and KP.3.1.1, XEC, and LP.8.1 currently circulating worldwide [2]. XEC, a recombination-derived lineage [3, 4] has acquired 2 new mutations (T22N, F59S), compared with the previously dominant KP.3 [3, 5]. Recently, XEC is increasingly outcompeted by LP.8.1, a KP.1.1.3 subvariant containing 9 new spike mutations versus JN.1 [4, 5] and 6 versus KP.2 [2]. LP.8.1 is increasingly prevalent globally, having become dominant in the United States [4, 6]. While LP.8.1 seems to be no more immune-evasive than XEC, it shows enhanced spike-hACE2 engagement, likely facilitating its gradual replacement over XEC [5]. Due to increasing global prevalence, both have been classified as variants under monitoring by the World Health Organization [2], accounting for 76% of sequenced variants (LP.8.1: 70%; XEC: 6%) in the US as of May 2025 [6].

Antigenic evolution within the JN.1 lineage led to authorization of monovalent JN.1- and KP.2-encoding mRNA COVID-19 vaccines (2024-2025 formula) [7, 8]. Preclinical versions of these vaccines evaluated in naive (2-dose primary series) and previously immunized mice (booster) were shown to neutralize vaccine-matched variants and cross-neutralize JN.1 subvariants KP.2, KP.3, LA.2, and XEC (XEC in naïve mice only) [9], supporting variant selection and updated vaccine approvals. The JN.1- and KP.2-encoding mRNA-1273 vaccines are expected to remain cross-reactive to emerging JN.1 lineage variants [4, 9]; however, clinical data on potential cross-neutralization are not yet available.

This study was conducted as part of an ongoing trial (NCT06585241) aiming to generate human serology data to assess immunogenicity and cross-neutralization of updated variant formulations of mRNA-1273 as new SARS-CoV-2 variants emerge. We evaluated the immunogenicity and cross-neutralization of the JN.1- encoding vaccine (mRNA-1273.167) and the KP.2-encoding vaccine (mRNA-1273.712) against vaccine-matched variants and currently circulating variants (KP.3.1.1, XEC, LP.8.1) [2, 6] in adults with previous COVID-19 mRNA vaccination; safety was also evaluated.

## Methods

### Study Design and Participants

This is an ongoing, open-label, single-arm, phase 3b/4 study evaluating immunogenicity and safety of monovalent formulations of mRNA-1273 encoding the SARS-CoV-2 Omicron variants JN.1 (mRNA-1273.167) and KP.2 (mRNA-1273.712). Eligible participants were US adults (≥18 years) with ≥3 previous mRNA COVID-19 vaccinations (≥2 mRNA COVID-19 vaccines and an XBB.1.5-encoding mRNA COVID-19 vaccine received between September 2023-August 2024). Participants received a 0.5-mL dose of a single intramuscular injection (50 µg) of either mRNA-1273.167 (Subprotocol 1) or mRNA-1273.712 (Subprotocol 2) and were followed for 1 month. Adults with known history of SARS-CoV-2 infection within 3 months before enrollment were excluded (**Supplementary Appendix**). The study is being conducted in accordance with the Declaration of Helsinki and Council for International Organizations of Medical Sciences international ethical guidelines, International Council for Harmonisation good clinical practice guidelines, and applicable laws and regulations.

### Patient Consent Statement

Protocol, informed consent form, and other relevant documents were reviewed and approved by the institutional review board/independent ethics committee (Advarra, Columbia, MD, USA) before study initiation. Informed consent was obtained prior to enrollment into the study.

### Study Vaccines

mRNA-1273.167 (Spikevax, Moderna, Inc.) contains 50 µg of mRNA encoding the full-length SARS-CoV-2 spike protein of the JN.1 variant with 2 proline residue substitutions to stabilize the spike protein into a prefusion conformation. mRNA-1273.712 contains 50 µg of mRNA encoding the prefusion-stabilized spike protein of the KP.2 variant.

### Immunogenicity and Safety Assessments

Sera for assessments of neutralizing antibody (nAb) responses were collected before vaccine administration (Day 1) and 4 weeks after immunization (Day 29). The nAb titers were quantified using a lentivirus-based SARS-CoV-2 pseudovirus neutralization assay (Duke University), described previously [10]. Safety assessments included adverse events (AEs) leading to study withdrawal, serious AEs (SAEs), and AEs of special interest (AESIs) from Day 1 through the end of the study.

### Statistical Analysis

Immunogenicity was evaluated in the per-protocol immunogenicity set (PPIS), comprising all participants who received a planned study vaccine, had a negative reverse transcriptase polymerase chain reaction (RT-PCR) test at Days 1 and 29, and no major protocol deviations impacting the key data. The primary endpoint was geometric mean (GM) titer and GM fold rise (GMFR) of nAb at Day 29 relative to baseline, with corresponding 95% confidence intervals. Safety was assessed in the safety set (all enrolled participants who received study intervention). No statistical hypothesis testing was performed; a descriptive summary of the primary immunogenicity objective was provided.

## Results

Overall, 50 participants were independently enrolled in each subprotocol and received mRNA-1273.167 (Subprotocol 1; median age, 63 years; 62% female; 86% Black/African American) or mRNA-1273.712 (Subprotocol 2; median age, 54 years; 66% female; 70% Black/African American). Median time on study was 30 days (mRNA-1273.167 cohort) and 29 days (mRNA-1273.712 cohort). Prior COVID-19 vaccination was categorized into 2, 3, 4, and ≥5 doses, with 76-80% of participants having received 3 doses across subprotocols (median time [interquartile range, IQR] since last dose: mRNA-1273.167, 310 days [303-316]; mRNA-1273.712, 330 days [277-338]). Prior COVID-19 vaccination included an original mRNA monovalent formulation (mRNA-1273 or BNT162b2: 100% recipients [mRNA-1273.167]; 96% recipients [mRNA-1273.712]), and Omicron bivalent formulation (mRNA-1273 or BNT162b2: 100% recipients [both subprotocols]). Baseline SARS-CoV-2 status was determined by virologic (RT-PCR) and/or serologic (anti-nucleocapsid binding antibody) evidence of SARS-CoV-2 infection on/before Day 1. No participants reported a known SARS-CoV-2 infection within the past 3 months. In Subprotocol 1, 39 (78.0%) participants were SARS-CoV-2 positive at baseline; PPIS included 48 (96.0%) participants (two were excluded due to a missing or positive SARS CoV-2 RT-PCR result on Day 29). In Subprotocol 2, 43 (86.0%) participants were baseline SARS-CoV-2 positive; PPIS included 49 (98.0%) participants (one was excluded due to a positive or missing SARS CoV-2 RT-PCR result on Day 29).

Both JN.1-encoding (mRNA-1273.167) and KP.2-encoding (mRNA-1273.712) vaccines elicited robust increases in nAb responses at Day 29 relative to baseline against matched variants (GMFR, 11.6-11.7) and cross-neutralized JN.1-lineage subvariants not matched to the vaccine (GMFR, mRNA-1273.167: KP.2, 8.1; KP.3.1.1, 10.5; XEC, 12.9; LP.8.1, 10.8; GMFR, mRNA-1273.712: JN.1, 10.8; KP.3.1.1, 12.4; XEC, 12.1; LP.8.1, 14.3; **Figure 1**). The highest titers were measured against JN.1 for both vaccines (GMT: mRNA-1273.167, 1670.0; mRNA-1273.712, 2796.4). Relative to the JN.1 reference, a reduction in cross-neutralization was observed against JN.1-lineage subvariants (KP.3.1.1, XEC, LP.8.1) for both vaccines, with the greatest reduction measured against KP.3.1.1 (**Figure 2**). Higher nAb responses were elicited by mRNA-1273.712 than mRNA-1273.167 against all tested variants (**Figure 2**).

**Figure 1.**
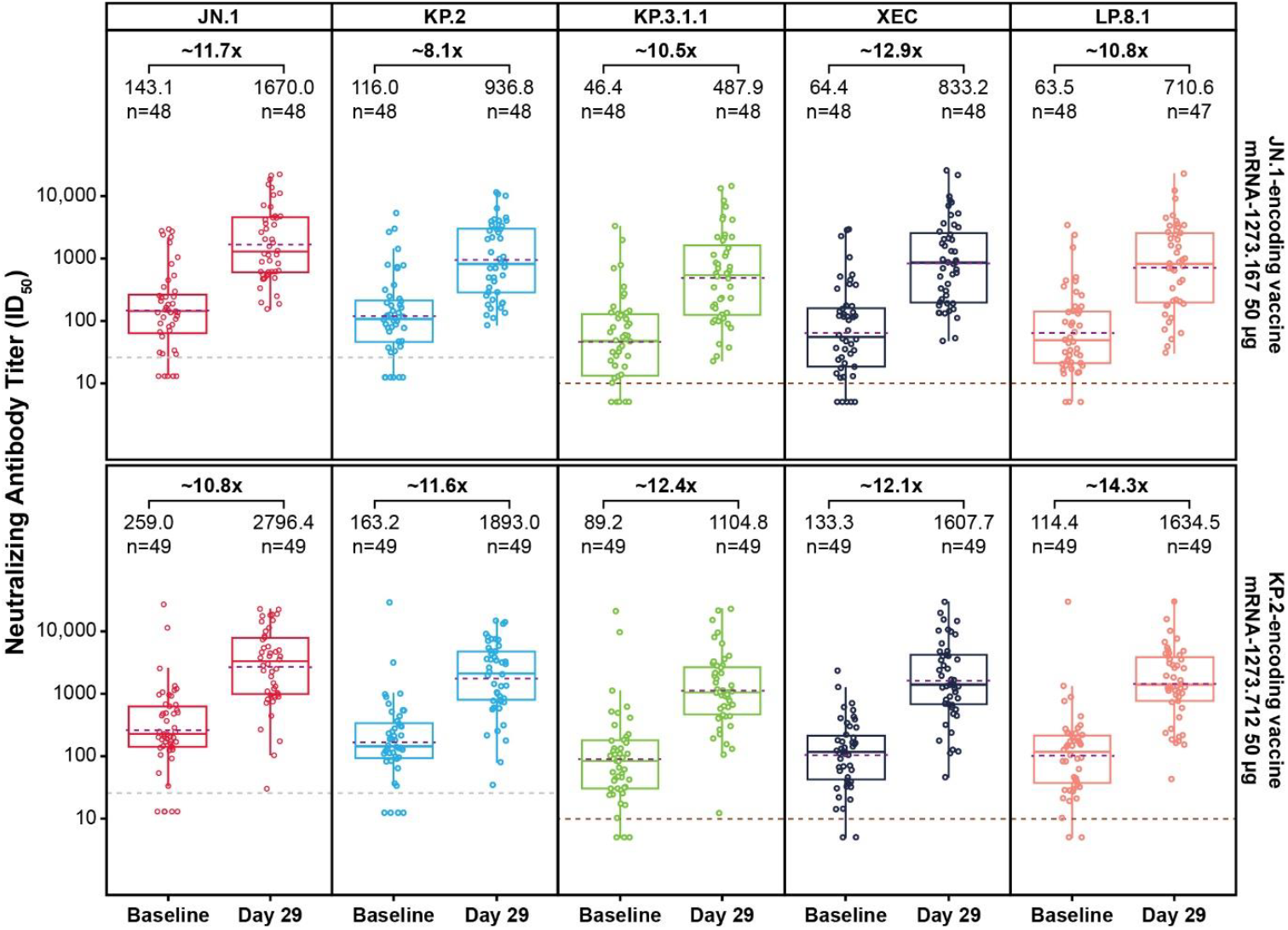
Neutralizing antibody responses (GMT and GMFR) elicited by mRNA- 1273.167 (JN.1-encoding vaccine) and mRNA-1273.712 (KP.2-encoding vaccine) against the vaccine-matched variants (JN.1, KP.2) and newly emerged subvariants (KP.3.1.1, XEC, LP.8.1), per-protocol immunogenicity set. The GMT (Day 1 and Day 29) with the corresponding GMFR (Day 29 relative to Day 1) are shown. The per-protocol immunogenicity set included participants who received the planned study intervention, had negative RT-PCR tests at baseline (Day 1) 1) and Day 29, and had no major protocol deviations that impacted the key data. The boundary of boxes represents the 25th (bottom) and 75th (top) percentiles of the GMT. The solid line inside the box represents the median (50th percentile) of the GMT. The dashed line inside the box represents the GMT. Whiskers (vertical lines) represent the lowest and highest data points within 1.5 of the interquartile range. The LLOQ is presented using a grey dashed line. The LOD is presented using a brown dashed line. Abbreviations: GMFR, geometric mean fold rise; GMT, geometric mean titer; ID50, 50% inhibitory dilution; LLOQ, lower limit of quantification; LOD, limit of detection; RT-PCR, reverse transcription polymerase chain reaction.

**Figure 2.**
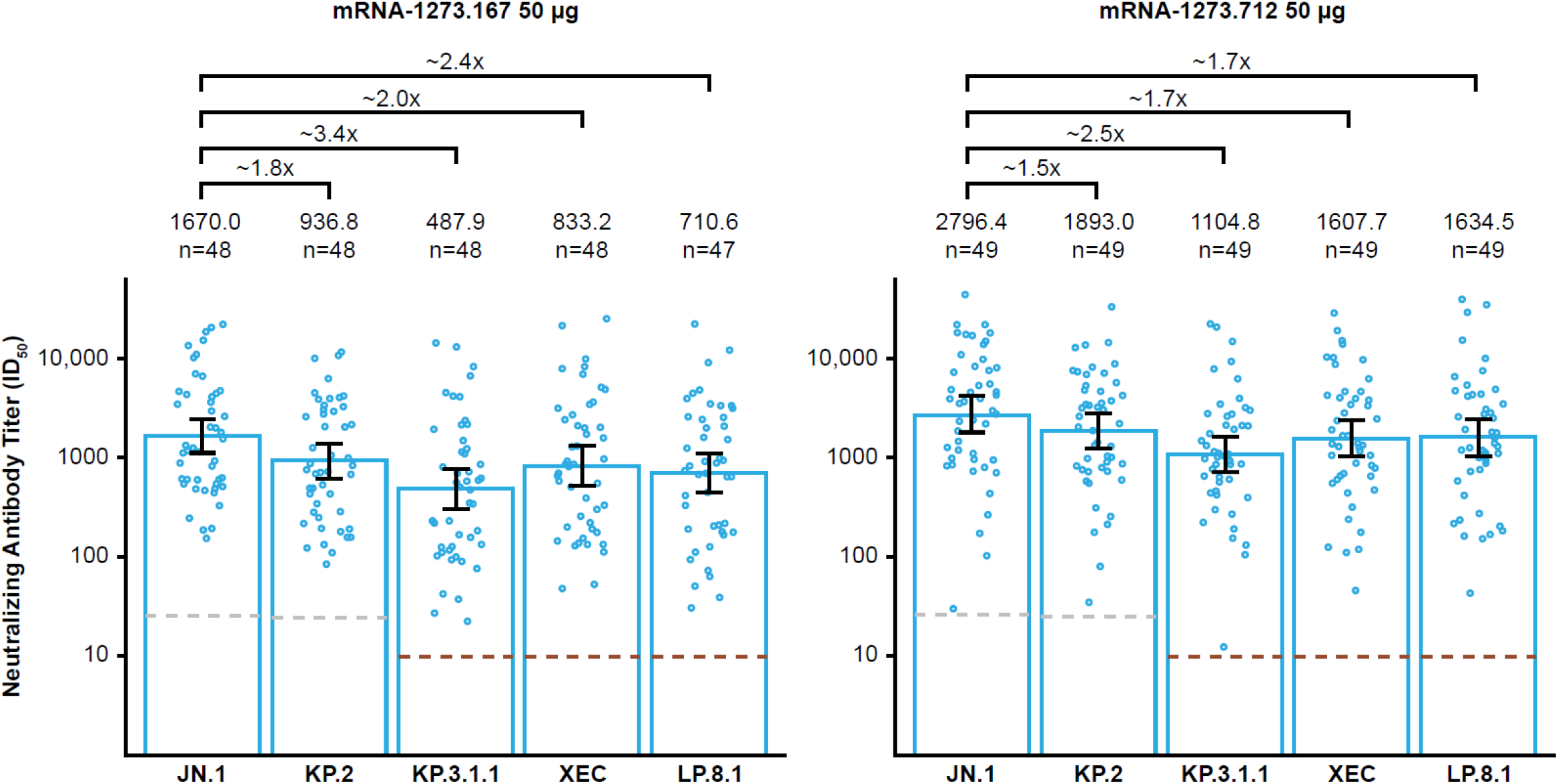
Geometric mean fold-drop in variant-specific neutralizing antibody responses at Day 29 after mRNA-1273.167 (JN.1-encoding vaccine) and mRNA- 1273.712 (KP.2-encoding vaccine), per-protocol immunogenicity set. The LLOQ is presented using a grey dashed line. The LOD is presented using a brown dashed line. Annotated fold change indicates fold-drop compared with the reference variant JN.1. Abbreviations: LLOQ, lower limit of quantification; LOD, limit of detection.

No SAEs, deaths, or AEs leading to study withdrawal were reported throughout the study. One AESI (pulmonary embolism) was reported in the study, which was considered unrelated to vaccination by the investigator. At 23 days post-vaccination in the JN.1 cohort, a participant in their 50s with hypothyroidism presented to the emergency department with chest pain/diaphoresis; a small pulmonary embolism was found, and the participant was treated with anticoagulants and then discharged on oral anticoagulants.

## Discussion

In this phase 3b/4 study evaluating adults with previous COVID-19 mRNA vaccination, monovalent JN.1-and KP.2-encoding mRNA-1273 vaccines (mRNA-1273.167, mRNA-1273.712) induced robust nAb responses against matched variants and cross-neutralized newly-emerged JN.1 subvariants (KP.3.1.1, XEC, LP.8.1). Cross-neutralization was reduced across all subvariants versus JN.1. No safety concerns were identified over the 1-month follow-up period. While both vaccines induced cross-neutralization consistent with the expected potent cross-reactivity of the JN.1- and KP.2-encoding vaccines against emerging JN.1-lineage subvariants [4, 9], decreased cross-neutralization measured against KP.3.1.1, XEC, and LP.8.1 for both vaccines suggested that recently emerged variants have developed some immune escape from responses induced by currently approved COVID-19 vaccine compositions. At Day 29, the KP.2-encoding vaccine (mRNA-1273.712) induced higher nAb responses than the JN.1-encoding vaccine (mRNA-1273.167) against all variants tested, suggesting an advantage in preventing escape. The highest reduction in cross-neutralization was observed for KP.3.1.1, consistent with data reporting increased resistance to serum neutralization of KP.3.1.1 versus XEC in KP.2 vaccine recipients [11]. The drop in cross-neutralization versus the JN.1 reference ranged between 1.5 and 3.4 after the JN.1- and KP.2-encoding vaccines, which was lower than that reported against JN.1 (∼5.8-fold reduction) after booster vaccination with the XBB.1.5-encoding mRNA-1273 vaccine authorized for the 2023-2024 season [10]. These data are consistent with the reported cross-neutralization of KP.2 monovalent boosters against JN.1 subvariants, including KP.3.1.1 and XEC, and with observations of reduced neutralization against these subvariants versus JN.1 or KP.2 [12, 13]. Similar findings were reported for the JN.1-encoding mRNA booster among healthcare workers, with the highest neutralization measured against JN.1, and 1.9- and 2.9-fold lower titers measured against KP.3.1.1 and XEC, respectively [14].

Study limitations include small sample size and lack of durability assessments of nAb responses. Higher baseline titers in the KP.2 cohort might have influenced higher nAb titers and fold-rises at Day 29, although greater fold-rises for KP.2, KP.3.1.1, and LP.8.1 were observed in the KP.2 cohort despite higher baseline titers. Participant enrollment in the JN.1 and KP.2 vaccine arms was non-randomized, and comparisons should be made cautiously.

In conclusion, the JN.1- and KP.2-encoding monovalent mRNA-1273 vaccines elicited robust nAb responses against matched variants and cross-neutralized currently circulating JN.1 subvariants (KP.3.1.1, XEC, LP.8.1), with reduced cross-neutralization observed against all subvariants tested. No safety concerns were identified.

## Data Availability

As the trial is ongoing, access to patient-level data presented in the article and supporting clinical documents by qualified external researchers who provide methodologically sound scientific proposals may be available upon reasonable request for products or indications that have been approved by regulators in the relevant markets and subject to review from 24 months after study completion. Such requests can be made to Moderna, Inc., 325 Binney Street, Cambridge, MA, 02142 USA < >. A materials transfer and/or data access agreement with the sponsor will be required for accessing shared data. All other relevant data are presented in the paper.

## Acknowledgments

Medical writing and editorial assistance were provided by Anja Varjačić, PhD, of MEDiSTRAVA in accordance with Good Publication Practice (GPP 2022) guidelines, funded by Moderna, Inc., and under the direction of the authors. These data were previously presented at the Congress of the European Society of Clinical Microbiology and Infectious Diseases (ESCMID Global), April 11-15, 2025, Vienna, Austria.

## Author contributions

A.L.F., B.G., D.K.E., A.N., S.H., X.C., E.d.W., V.U., F.P., R.D., and S.C. conceived and planned the study. A.L.F., B.G., A.N., K.J., S.H., E.d.W., V.U., R.D., and D.C.M. collected the data. A.L.F., B.G., D.K.E., A.N., S.H., X.C., E.d.W., V.U., R.D., D.C.M., and S.C. contributed to data analysis and interpretation. All authors contributed to drafting of the manuscript and contributed to the critical revision of the manuscript for important intellectual content. All authors met the authorship criteria and approved the publication.

## Figure Alt Text

Figure 1: Graphs showing neutralizing antibody responses against vaccine-matched variants and newly emerged variants at baseline and Day 29 after JN.1- and KP.2- encoding mRNA-1273.712 vaccines, illustrated with boxplots and individual data points.

Figure 2: Graphs comparing geometric mean fold-drop in variant-specific neutralizing antibody responses at Day 29 after JN.1- and KP.2-encoding mRNA- 1273.712 vaccines, illustrated with bars and individual data points.

