## Supplementary Appendix for "Immunogenicity of JN.1- and KP.2-Encoding mRNA COVID-19 Vaccines Against JN.1 Subvariants in Adult Participants"

*Inclusion Criteria*

Participants were eligible for inclusion in the study if all the following criteria were met:

1. Male or female; ≥18 years of age at the time of signing the informed consent.
2. Participants had previous mRNA COVID-19 vaccination (≥2 mRNA COVID-19 vaccines and an XBB.1.5-encoding mRNA COVID-19 variant-encoding vaccine received during the period between September 2023-August 2024).
3. Participant understood and was willing and physically able to comply with protocol-mandated follow-up, including all procedures, by investigator's assessment.
4. Female participants of childbearing potential were eligible for enrollment in the study if all the following criteria were fulfilled:
    a. Had a negative pregnancy test at the screening visit and on the day of vaccination (Day 1) prior to the administration of vaccine dose.
    b. Had practiced adequate contraception or had abstained from all activities that could have resulted in pregnancy for ≥28 days prior to the first dose (Day 1). Adequate female contraception was defined as consistent and correct use of a local health authority approved contraceptive method in accordance with the product label.
    c. Had agreed to continue adequate contraception through 28 days following vaccine administration.

       *Note*: Contraceptive use should have been consistent with local regulations regarding the methods of contraception for those participating in clinical studies.

5. Capable of giving signed informed consent, which included compliance with the requirements and restrictions listed in the informed consent form and in the protocol.

*Exclusion Criteria*

Participants were excluded from the study if any of the following criteria applied:

1. History of SARS-CoV-2 infection within 3 months prior to enrollment.
2. Was acutely ill or febrile (temperature ≥38.0 °C [≥100.4°F]) less than 72 hours prior to or at the screening visit or Day 1. Participants meeting this criterion could be rescheduled within the screening window and would have retained their initially assigned participant number.
3. History of a diagnosis or condition that, by the judgment of the investigator, was clinically unstable, or might have affected participant safety, assessment of study endpoints and immune response, or adherence to study procedures.
    - Clinically unstable was defined as a diagnosis or condition requiring changes in management or medication within 60 days prior to screening and included ongoing workup of an undiagnosed illness that could lead to a new diagnosis or condition.
    - Asymptomatic conditions and conditions with no evidence of end organ involvement (eg, mild hypertension, dyslipidemia) were not exclusionary, provided that they were appropriately managed and clinically stable (ie, unlikely to result in symptomatic illness within the time course of study). Illnesses or conditions could be exclusionary, even if otherwise stable, due to therapies used to treat them (eg, immunosuppressive treatments), at the discretion of the investigator.

4. History of anaphylaxis or severe hypersensitivity reaction requiring medical intervention after receipt of any mRNA vaccine or therapeutic or any components of an mRNA vaccine or therapeutic.
5. History of myocarditis, pericarditis, or myopericarditis within 90 days prior to the screening visit.
    - Participants who had not returned to baseline after their convalescent period would also have been excluded.
6. History of Guillain-Barré syndrome.
7. History of coagulopathy or bleeding disorder that was considered a contraindication to intramuscular injection or phlebotomy.
8. Reported history of congenital or acquired immunodeficiency (eg, HIV), immunosuppressive condition, asplenia, or recurrent severe infections.
9. Any medical, psychiatric, or occupational condition, including reported history of drug or alcohol abuse, that, in the opinion of the Investigator, might have posed additional risk due to participation in the study or could have interfered with the interpretation of study results.
10. Receipt of the following:
    a. COVID-19 vaccine within 3 months prior to enrollment.
    b. Any licensed non-COVID-19 vaccine within 28 days before or planned receipt within 28 days after the study intervention, except an influenza vaccine, which might have been given 14 days before or after receipt of study intervention.
    c. Systemic immunosuppressants or immune-modifying drugs for >14 days total, within 6 months prior to screening (for corticosteroids ≥10 mg/day of prednisone equivalent) or was anticipating the need for

immunosuppressive treatment at any time during participation in the study. Participants who received epidural or major joint intra-articular injections (eg, knee, hip, shoulder) within 28 days prior to or within 28 days after Day 1 were excluded.

*Note*: Inhaled, nasal, and topical steroids were allowed.

d. Systemic immunoglobulins or blood products within 3 months prior to the screening visit (Day 0) or planned for receipt during the study.

11. Had donated ≥450 mL of blood products within 28 days prior to the screening visit or planned to donate blood products during the study.

12. Had participated in an interventional clinical study within 28 days prior to the screening visit or planned to participate in an interventional clinical study of an investigational vaccine or drug while participating in this study.

    *Note*: Interventions such as counseling, biofeedback, and cognitive therapy were not exclusionary.

13. Was working or had worked as study personnel or was an immediate family member or house member of study personnel, study site staff, or Sponsor personnel.
